# Unifying diagnostic criteria for gestational diabetes mellitus

**DOI:** 10.1101/2021.07.01.21259865

**Authors:** Suhail A. R. Doi, Mohammed Bashir, Michael T. Sheehan, Adedayo A. Onitilo, Tawanda Chivese, Ibrahim M. Ibrahim, Stephen F. Beer, Luis Furuya-Kanamori, Abdul-Badi Abou-Samra, H. David McIntyre

## Abstract

**AIMS:** Disagreement about the appropriate criteria for the diagnosis of gestational diabetes mellitus (GDM) persists. This study examines an alternative approach which combines information from all time-points on the glucose tolerance test (GTT) into a single index and expands the GDM spectrum into four categories using data from three geographically and ethnically distinct populations.

**METHODS:** A retrospective observational study design was used. Data from Wisconsin, USA (723 women) was used in derivation of the criterion and data from Doha, Qatar (1284 women) and Cape Town, South Africa (220 women) for confirmation. Pregnant women without pre-existing diabetes with a GTT done between 23 and 30 weeks gestation were included. A novel index was derived from the GTT termed the weighted average glucose (wAG). This was categorized into four pre-defined groups (henceforth National Priorities Research Program (NPRP) criterion); i) normal gestational glycemia (NGG), ii) impaired gestational glycemia (IGG), iii) GDM and iv) high risk GDM (hGDM).

**RESULTS:** In the Doha cohort, compared to the NGG group, the odds of large for gestational age babies increased 1.33 fold (P=0.432), 2.86 fold (P<0.001) and 3.35 fold (P<0.001) in the IGG, GDM and hGDM groups respectively. The odds of pregnancy induced hypertension increased 2.10 fold (P=0.024) in GDM & hGDM groups compared to the IGG and NGG groups. In the Cape Town cohort, a third of women in the GDM group and two-thirds in the hGDM group progressed to T2DM at 5 years.

**CONCLUSIONS:** The NPRP categorization identifies four distinct risk clusters of glycemia in pregnancy which may aid better decision making in routine management, avoid potential over-diagnosis of women at lower risk of complications and assist with diabetes prevention in women at high-risk after an index pregnancy with GDM.

## 1. INTRODUCTION

Gestational diabetes mellitus (GDM) is defined as hyperglycaemia first detected during pregnancy that is neither type 1 diabetes mellitus (T1DM) or type 2 diabetes mellitus (T2DM) [1,2]. GDM is associated with an increased risk of maternal and fetal complications, increased medical cost [3] and can only be established through biochemical testing, typically between 24-28 weeks’ gestation. Despite wide agreement that any form of hyperglycaemia should be managed during pregnancy, there is still a substantial disagreement on which diagnostic process should be used and which glucose threshold(s) merit a diagnosis of GDM [4]. The two widely used criteria for the diagnosis of GDM rely on individual time-point thresholds on a 2-hour [5] or a 3-hour [6] glucose tolerance test (GTT), and require elevated values at one or two time points respectively for the diagnosis. As such, the number of elevated time-point values required and different glycaemic thresholds will alter the prevalence of GDM [7]. The changes in prevalence have, nevertheless, not been associated with maternal or perinatal benefit overall [8] implying that [9] broadening the diagnosis through use of the IADPSG single-step approach (over the two step approach) remain insufficient to justify the increased costs [9].

The increase in prevalence from application of the International Association for the Diabetes in Pregnancy Study Group (IADPSG) 2010 [5] definition of GDM in Denmark has been reported to result in a marked increase in GDM prevalence to 40% without convincing evidence of adverse pregnancy outcomes [10]. Although this could be specific to Denmark, it raises serious questions about uniform application of current GDM diagnostic thresholds across the world, especially for criteria based on individual cglucose thresholds on the GTT. This is important given that 75-85% of the women diagnosed with GDM do not require any pharmacological treatment [11–14], as it can be argued that the current GDM diagnostic thresholds over-diagnose women at lower risk of complications. The latter is important since women at low risk may not benefit from reduced adverse pregnancy outcomes with treatment, but still experience higher induction rates, Caesarean section rates and lower gestational age at birth [15] with their consequent long-term risks, anxiety and economic burden [8,16–18]. Additionally, GDM is a risk factor for T2DM [19], and there is therefore a need to refocus attention on criteria that distinguish women at higher risk of such progression [8]. Current criteria may also not address the latter optimally since progression to T2DM also varies with the type and number of abnormal GTT values [20].

In an effort to address these problems, one suggestion has been adjustment of fasting venous plasma glucose (FVPG) concentrations to the local context through standard deviation based z-score transformation [21]. However, these z-scores are sensitive to the population distribution of normal and abnormal values and, unless based on a large representative sample, can make the cutoffs difficult to justify. In addition, diabetes outside pregnancy is defined worldwide by fixed glucose levels and GDM should arguably be no different. It is therefore unlikely that reducing single thresholds will suffice [10,22]. What is needed therefore is a criterion based on the whole GTT in lieu of criteria based on individual time-points on the GTT, is not population specific and identifies women and babies at high risk and assigns a GDM diagnosis label that does no harm to women at lower risk [23]. This study aims to address these problems with GDM diagnosis through examination of data from three continents, assessing the utility of combining information from all time-points on the GTT and expanding the GDM spectrum beyond the current binary approach.

## 2. MATERIALS AND METHODS

Data was collected through the electronic medical record (EMR) at the Marshfield Clinic in Wisconsin and the Hamad General Hospital, Hamad Medical Corporation, in Doha, while the South African Data was prospectively collected. Given the retrospective, de-identified nature of data collection, the institutional review board at the Marshfield Clinic approved this study as minimal risk while the other two dataset did not require approval having been previously published with relevant approvals. The methods and reporting of results adhere to the STROBE (STrengthening the Reporting of OBservational studies in Epidemiology) guidelines. Women were identified using the laboratory database. Exclusions were: women who did not complete the full two hour GTT; women with a FPG ≥ 7.0 mmol/l and/or 2 h PG ≥ 11.1 mmol/l who were deemed to have T2DM. All women thus identified were managed by local protocols at relevant institutions.

### 2.1 Data sources

At Wisconsin, the target population was mothers who underwent a GTT (after being identified by a ‘first step’ universal 50g random glucose challenge test (GCT) with a value at 1h ≥7.5 mmol/L) and gave birth between November 2014 and November 2019. GDM was defined according to the criteria of Carpenter and Coustan [6] (fasting plasma glucose (FPG) >5.2 mmol/l, >10 mmol/l at 1 h, >8.6 mmol/l at 2 h, or >7.7 mmol/l at 3 h) with two or more plasma glucose levels exceeding these cutoff values required for the diagnosis.

At Doha, the data used has previously been reported [7] and was a convenience sample that included all women who underwent a 75 g OGTT between January 2016 and April 2016 with and without GDM at a tertiary hospital of the Hamad Medical Corporation (Women’s Wellness and Research Center) in Qatar. This is the main maternity hospital in Qatar with 16,000–18,000 births annually. GDM was defined in this population using the WHO2013/IADPSG criteria [5,24].

The Cape Town data is from the *PROgression to type 2 Diabetes* (PRO2D) study which investigated the progression to T2DM 5 years after a GDM pregnancy in Cape Town, South Africa. In brief, all women diagnosed with GDM at a major tertiary referral hospital in Cape Town, South Africa, between 1 August 2010 and 30 September 2011, were recalled for diabetes assessment 5-6 years after the index pregnancy. Women were diagnosed using the revised Western Cape guideline that was based on literature reviewed till June 2010 and expert consensus (75g 2-hr GTT with either a fasting value >5.5 mmol/L or a 2-hour value ≥7.8 mmol/L) [25], From the 1st of May 2016 till the 30th March 2017, consecutively recruited women were recalled and assessed for diabetes using a standard 2-hour GTT, and HbA1c. The detailed description of the PRO2D study has been published elsewhere [25,26].

### 2.2 Variables of interest and measurements

Four categories of variables were extracted from the electronic medical record: *demographic information*; *risk factors; test result & timing-related*; and *outcomes*. Large for gestational age (LGA; >90^th^ percentile), small for gestational age (SGA; <10^th^ percentile) and appropriate for gestational age (AGA)) were calculated using the Fenton growth calculator [27,28]. Lists of variables extracted and descriptive analyses are reported in the supplementary material. Participants underwent a standard GTT, with the use of either a 100-g (USA) or 75-g (Qatar, South Africa) oral glucose load, between 23 and 30 weeks of gestation (target time of testing, 24-28 weeks). GTTs done before 23 weeks and after 30 weeks were not included in the analysis. Details of glucose measurement methods and the difference between plasma glucose levels on the 100g and 75g GTT are detailed in the supplementary material.

### 2.3 Statistical Analysis

All GTT glucose values were converted to SI units (mmol/L) for analysis. A principal components analysis (PCA) was carried out using the Wisconsin dataset to derive the criterion using the following variables: z-score for GTT glucose 0h, z-score for GTT glucose 1h, z-score for GTT glucose 2h, z-score for GTT glucose 3h, GDM status (ICD-10: O24.4), GDM status (NDDG criterion [29]), GDM status (Carpenter-Coustan [6]), GDM status (IADPSG [5]), gestational age at birth (weeks), birth weight (grams) and LGA status. Two components were extracted, and it was expected that the first component would load variables on the GDM construct and the second on the fetal size construct. As expected, the last three variables loaded predominantly on the fetal size component. The rotated PCA loadings, which are the covariances/correlations between the original variables and the unit-scaled GDM component were taken as weights for each of the first three GTT glucose points and rescaled to sum to 1.

The GTT 0h, 1h and 2h glucose values were then combined into a weighted average glucose (wAG) using these individual weights. Based on the fact that GDM is a reflection of both impaired beta-cell function and increasing insulin resistance in pregnancy, the women were classified into groups based on the optimal cutoff points for wAG based on the median for two variables: history of previous GDM and pre-pregnancy obesity. This was justified given recent data that suggest that the latter variables are individually significantly associated (about twice the odds) with need for insulin therapy, and by extension, with metabolic severity [30]. This gave three cutoffs (median wAG with both absent, either present or both present). The four groups created by the three cut-offs were pre-defined as i) normal gestational glycemia (NGG), ii) impaired gestational glycemia (IGG), iii) GDM and iv) high risk GDM (hGDM) respectively. These labels were chosen as concordant with metabolic severity since this corresponds to insulin resistance since the latter groupings correlate well with changes in mean glucose levels on the GTT as well as with pregnancy outcomes [31]. These four groups based on the wAG cutoffs, define the National Priorities Research Program (NPRP) GDM criterion.

Next, the NPRP criterion was applied to the Doha data for confirmation and the performance vis-a-vis the NPRP criterion and clinical parameters were reported. Given that clinical action is usually prompted by the diagnosis of GDM, the analysis also looked at the subset of women with the GDM diagnosis in terms of the concordance of the treatments used with the NPRP categories.

Finally, the NPRP criterion was applied to the South African GDM data, to assess whether this criterion aligned with future T2DM at 5 years follow-up in the cohort of women with previous GDM. Both the risk category based on Kohler at baseline and the occurrence of T2DM at 5 year follow-up were compared across NPRP categories.

All analyses were conducted using Stata 15 MP4 (StataCorp, College Station, TX, USA). All increase or decrease in risk statements refer to the estimated odds ratio (OR) and not the P values. The P values only indicate degree of evidence against the model hypothesis (i.e. OR=1) at the current sample size. Thus, for P values above 0.05, the interpretation regarding the estimated effect of treatment does not change, only the degree of evidence against the model hypothesis changes. Exact P values were reported throughout.

## 3. RESULTS

### 3.1 Criterion Determination (USA, Wisconsin Data)

The weights for GTT 0h, 1h and 2h values were 0.28, 0.36 and 0.36 respectively. The wAG for each participant was computed as follows:

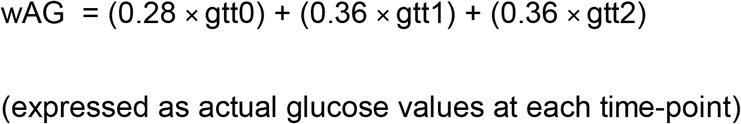

There were 723 participants with a GTT done between 23 and 30 weeks of gestation and the wAG could be calculated for 697 women. The range of wAG was 4.2 to 11.9 mmol/L (median 7.2 mmol/L). The empirical wAG cutoffs were 6.8, 7.5 and 8.6 mmol/L and were used to classify the women into the four predefined GDM risk groups (Table 1). Of the 697 women, 257 (36.9%), 159 (22.8%), 196 (28.1%) and 85 (12.2%) were classified as NGG, IGG, GDM and hGDM respectively. Descriptive analyses are reported in Table S1 (supplementary material).

**Table 1.** Gestational diabetes NPRP risk group classification

| **GDM category** | **wAG threshold**<br>**(mmol/L)** |
| --- | --- |
| **Normal gestational glycemia** | $\leq 6.8$ |
| **Impaired gestational glycemia** | $>6.8 \leq 7.5$ |
| **Gestational diabetes** | $>7.5 \leq 8.6$ |
| **High risk gestational diabetes** | $> 8.6$ |

\*wAG; weighted average glucose

#### 3.1.1 Comparisons with the NGG category

The odds of a history of previous GDM was 2.22 fold higher (P=0.148) in IGG, 8.16 fold higher (P<0.001) in GDM and 26.55 fold higher (P<0.001) in the hGDM categories respectively. The odds of pre-pregnancy obesity was 1.93 fold higher (P=0.009) in IGG, 2.67 fold higher (P<0.001) in GDM and 4.24 fold higher (P<0.001) in the hGDM categories respectively. This suggested a dose dependent relationship in both cases which was expected given these were used to define the cutoffs. In addition, the odds of a LGA baby was 1.48 fold higher (P=0.269) in IGG, 2.15 fold higher (P=0.015) in GDM and 2.67 fold higher (P=0.009) in the hGDM categories respectively again consistent with a dose dependent relationship.

### 3.2 Criterion Assessment and Confirmation (Doha, Qatar)

There were 401 women diagnosed with GDM and 883 women not diagnosed with GDM. The wAG was calculable in 374 of 401 women with GDM, the range being 4.8 to 13.0 mmol/L (median 8.1 mmol/L). The wAG was calculable in 882 of 883 women not diagnosed with GDM, the range being 3.4 to 8.0 mmol/L (median 5.9 mmol/L). Of these 1256 women, 762 (60.7%), 194 (15.5%), 178 (14.2%) and 122 (9.7%) were classified as NGG, IGG, GDM and hGDM respectively. Descriptive analyses are reported in Table S2 (supplementary material).

#### 3.2.1 Metabolic outcomes

The average first HbA1c was 5.15% (32.8 mmol/mol) in the NGG group and *increased* by 0.20% (P=0.045), 0.26% (P=0.004) and by 0.36% (P<0.001) in IGG, GDM and hGDM groups respectively. Compared to the NGG group, the odds of obesity increased 2.56 fold (P<0.001) in IGG, 3.78 fold in GDM (P<0.001) and 4.63 fold in the hGDM group (P<0.001). A similar trend was noted for women who were overweight (OR 1.75, P=0.014; OR 2.75, P=<0.001; OR 2.42, P=0.007 respectively).

#### 3.2.2 Maternal and perinatal outcomes

The odds of LGA babies increased over the NGG group by 1.33 fold (P=0.432), 2.86 fold (P<0.001) and 3.35 fold (P<0.001) in the IGG, GDM and hGDM groups respectively. While LGA demonstrated a dose-response relationship with glycemic categories, the other pregnancy related outcomes were seen mainly in the two GDM groups and the odds of the newborn’s NICU admission (OR 1.46; P=0.071), cesarean delivery (OR 1.53; P=0.002), preterm (<37 weeks gestational age) delivery (OR 1.51; P=0.023), induction of labour (OR 1.74; P=0.003) and PIH (OR 2.10; P=0.024) were increased in the women in the GDM groups compared to the women in the other two groups, despite having been modulated by treatment. There was some evidence against the model hypothesis for excess gestational weight gain (OR 1.15; P=0.321) and stillbirth (OR 1.84; P=0.335), but almost no evidence against the model hypothesis for pre-eclampsia (OR 1.16; P=0.718) in GDM compared to the non-GDM groups at this sample size. Numbers are given in Table 2.

**Table 2.** Risk factors and outcomes by NPRP categories

| Risk factors and outcomes | NGG<br>(wAG ≤6.8<br>mmol/L) | IGG<br>(wAG >6.8 to<br>≤7.5 mmol/L) | GDM<br>(wAG >7.5 to<br>≤8.6 mmol/L) | hGDM<br>(wAG >8.6<br>mmol/L) |
| --- | --- | --- | --- | --- |
| UNITED STATES |  |  |  |  |
| History of previous GDM | 6/257 (2.3%) | 8/159 (5.0%) | 32/196 (16.3%) | 33/85 (38.8%) |
| Pre-pregnancy obesity | 46/178 (25.8%) | 49/122 (40.2%) | 68/141 (48.2%) | 34/57 (59.6%) |
| LGA baby | 19/222 (8.6%) | 12/135 (8.9%) | 29/173 (16.8%) | 15/75 (20.0%) |
| QATAR |  |  |  |  |
| First mean% (mmol/mol) | 5.15% (32.6) | 5.35% (35) | 5.41% (35.6) | 5.51% (36.7) |
| Obesity | 244/745 (32.8%) | 90/191 (47.1%) | 86/176 (48.9%) | 67/119 (56.3%) |
| LGA baby | 32/646 (5.0%) | 11/170 (6.5%) | 21/162 (13.0%) | 15/101 (14.9%) |
| On insulin (diagnosed GDM only) | 8/27 (29.6%) | 22/65 (33.9%) | 62/160 (38.8%) | 77/122 (63.1%) |
| On metformin (diagnosed GDM only) | 7/27 (25.9%) | 21/65 (32.3%) | 55/160 (34.4%) | 67/122 (54.9%) |
| PIH | 25/956 (2.6%) |  | 16/300 (5.3%) |  |
| NICU admission | 84/956 (8.8%) |  | 37/300 (12.3%) |  |
| Caesarean delivery | 354/956 (37.0%) |  | 142/300 (47.3%) |  |
| Preterm delivery | 119/954 (12.5%) |  | 53/300 (17.7%) |  |
| Induction of labour | 103/956 (10.8%) |  | 52/300 (17.3%) |  |
| SOUTH AFRICA |  |  |  |  |
| T2DM at 5 years<sup>a</sup> | 0/12 (0%) | 2/18 (11.1%) | 32/88 (36.4%) | 71/102 (69.6%) |
| Kohler risk score (IQR)<sup>b,c</sup><br>(N=214) | 213.9 (120.3,<br>295.7)<br>N=12 | 204.8 (166.3,<br>256.3)<br>N=17 | 191.2 (151.4,<br>275.6)<br>N=85 | 285.3 (204.0,<br>341.4)<br>N=100 |

<sup>a</sup> Pearson chi2(3) = 44.8, P<0.001
<sup>b</sup> Kruskal-Wallis equality-of-populations rank test, Chi2(3)=23.2; P<0.001
<sup>c</sup> Kruskal-Wallis equality-of-populations rank test after exclusion of hGDM, Chi2(2)=0.02; P=0.990
Range of Kohler score is 54 – 380 with ≤140 (low), 141 – 220 (medium), 221 – 300 (high) and >300 very high risk respectively

#### 3.2.3 Treatment requirements by NPRP category

An analysis of treatment (Table 2) was undertaken amongst those with a diagnosis of GDM in this cohort. In this group, compared to those classified as hGDM, the odds of insulin treatment decreased by two-thirds in those classified as GDM (OR 0.37; P=<0.001) or IGG (OR 0.30; P<0.001) and decreased by three-quarters (OR 0.25; P=0.002) in those classified as NGG by the NPRP criterion. Similarly, compared to the hGDM group, the odds of metformin treatment decreased by two-thirds in GDM (OR=0.37; P<0.001) and by 90% in IGG (OR 0.10; P<0.001) and by more than 99% (OR 0.008; P<0.001) in the NGG group.

### 3.3 Future Diabetes Risk (Cape Town, South Africa)

Descriptive analyses are reported in Table S3 (supplementary material). Only women with GDM by local criteria were followed-up and there was a clear relationship between future diabetes at 5 years and the NPRP GDM status in pregnancy with risk increasing in the GDM and hGDM groups. A third of women in the GDM group and two-thirds in the hGDM group progressed to T2DM at 5 years (Table 2). Compared to the NGG/IGG group (2/30), the GDM group had a 8 fold (32/88) increase in odds (P=0.007) and hGDM group had a 32.1 fold (71/102) increase in odds (P<0.001) of T2DM at five years. When looking at the Kohler risk score [32] in relation to the GDM risk groups, the risk scores were only elevated in the hGDM group compared to the other groups which all had median risk scores at the medium risk level (Table 2), suggesting that the NPRP risk category takes precedence over the risk factors in terms of future T2DM.

## 4. DISCUSSION

In this study, we have developed and validated a wAG measure from the GTT and defined four distinct categories; NGG, IGG, GDM and hGDM. These four categories identify women who are at progressively higher risk of both immediate pregnancy-complications and subsequent development of T2DM. The populations used for the development and confirmation of the wAG are distinct both geographically and racially, and the results were consistent across three continents. Hence, we propose the use of the wAG based NPRP-criteria for the screening and classification of glucose tolerance during pregnancy.

PIH and LGA are the most common and consistently reported maternal and fetal complications of GDM, respectively [33]. The risk of these complications showed a clear dose-dependent relationship with the NPRP categories. There are also longer term consequences of GDM since the metabolic adaptations during pregnancy place additional stress on β-cells that are already defective pre-pregnancy. The progressive β-cell dysfunction after an index GDM pregnancy due to retention of excessive gestational weight gain and increases in insulin resistance worsens the risk of T2DM in the years after pregnancy [33] and was predicted, in a dose dependent fashion, by increasing NPRP categories at 5 years. In addition, GDM risk factors [32,33] considered within the Kohler score [32] also demonstrates a dose dependent relationship with the NPRP criterion.

Based on the HAPO study of maternal and fetal outcomes in 23,316 women [34], it was clear that the risk associated with hyperglycemia in pregnancy is a continuum with no ‘natural inflection points’. Therefore, women with equivalent levels of glycaemia-associated risk of adverse pregnancy outcomes were recommended to be grouped together by standardized threshold plasma glucose values. Despite this, only two groups were defined – normal and GDM [5,34], based on an adjusted odds ratio threshold of 1.75 (compared with the odds at the HAPO cohort mean) of delivering an infant affected by key fetal complications [35]. Our study extends this principle to a spectrum of GDM categories, thus defining risk status better. In the absence of any natural threshold, the precise numeric cutoff values of maternal glycaemia used for diagnosis as well as number of abnormal time-points have led to ongoing debate and may not also be suitable for uniform worldwide application [5,10,34,36–39]. This study suggests that the latter is likely a function of the binary classification and the single value cutoffs used by the IADPSG as we did not note a difference in performance of the NPRP criterion over three continents. Also, integrating three GTT glucose values simultaneously eliminates the need to decide on the use of one versus two abnormal thresholds and allows risk categories to be defined incrementally based on overall glucose exposure. Our study also addresses the call for a more flexible approach [40] without the need for differing diagnostic processes and glucose thresholds in specific geographic regions and ethnic groups.

This study raises some important practical points which merit further consideration and evaluation of limitations to improve clinical practice. First, we use retrospective data and therefore these findings will benefit from prospective evaluation. Second, we acknowledge that the four categories proposed in this paper could generate some uncertainty for both women and their care givers who are accustomed to the binary classification. This will require a new therapeutic approach based on variations in risk of short and long term outcomes. However, these categories could be instrumental in guiding care during pregnancy.

In conclusion, these new criteria reduce the challenge physicians face with identification of those who will require pharmacotherapy as the majority of the women will not. The new IGG category, despite being more obese than the the normal group, appear not to be at higher risk of pregnancy complications, have a reduced rate of need for pharmacotherapy during pregnancy and have only a modestly increased risk of progression to T2DM. The implication is therefore less intensive intervention, focusing primarily on nutritional therapy and advice on healthy physical activity [41] in this group. Similarly, GDM and hGDM categories might be started concurrently on pharmacotherapy [42] and nutritional therapy, and hGDM may be considered for insulin therapy sooner. Post-delivery, women with hGDM (and perhaps GDM if resources allow) should be the major target group for a coordinated diabetes prevention program including dietary and life-style intervention, Metformin, or both. As such, the four identified categories can provide more clarity and better resource allocation in the management of women at high-risk during and after pregnancy.

## Data Availability

Available from the authors on reasonable request

